# Complexity of hospital demand during the COVID-19 pandemic in Mexico City

**DOI:** 10.1101/2025.01.30.25321424

**Authors:** Guillermo de Anda-Jáuregui, José Sifuentes-Osornio, Ofelia Angulo-Guerrero, Juan L Díaz-De-León-Santiago, Héctor Benítez-Pérez, Luis A Herrera, Oliva López-Arellano, Arturo Revuelta-Herrera, Ana R Rosales-Tapia, Manuel Suárez-Lastra, David Kershenobich, Rosaura Ruiz-Gutiérrez, Enrique Hernández-Lemus

## Abstract

**Background:** The COVID-19 pandemic posed unprecedented challenges to healthcare systems worldwide. In densely populated urban areas such as Mexico City, the strain on hospitals was amplified due to the high volume of cases and resource limitations. Understanding the spatial and temporal dynamics of hospital demand is crucial for informing effective public health strategies and improving system resilience.

**Methods:** A retrospective analysis of COVID-19 hospitalization data in Mexico City was conducted utilizing a line-list dataset from the SISVER surveillance system. The analysis included the spatial distribution of hospital demand using the weighted centroid of hospitalizations as well as a model system of the interactions between residential areas and hospitals as a bipartite network. The emergence of giant components in the network was used as indicators of system strain and the relationship between network strain and patient outcomes was evaluated.

**Findings:** Hospital demand in Mexico City exhibited significant spatial dynamics, with a northward shift in the weighted centroid of hospitalizations as the pandemic progressed. Despite the changing distribution of cases, a small subset of 17 hospitals managed the majority of hospitalizations. During high-demand periods, the network transitioned to a more disordered state, characterized by a giant component encompassing multiple neighborhoods. This disordered strain was associated with higher case fatality rates, particularly in patients over 40 years of age.

**Interpretation:** Our findings highlight the complex, adaptive nature of the healthcare system in response to the pandemic. The emergence of giant components in the hospital demand network can serve as an early warning indicator of the health system overload. Adaptive measures, such as the establishment of temporary COVID-19 units, were effective in mitigating strain. These insights can guide future public health strategies for rapid response and resource allocation in similar crises.

**Funding:** This project was partially funded through CONACYT Project 320557 (to GA-J).

## Introduction

The COVID-19 pandemic brought unprecedented challenges to the social, public, and individual sectors. Aside from the obvious myriads of unfortunate events at the personal level, health systems were affected in an unparalleled manner, the major medical institutions were out of capacity, medical personnel were overwhelmed and resources became scarce (1–3). Furthermore the spatial and temporal patterns in which these phenomena occurred unveil underlying and structural constraints that are worth considering to revise, more so in the face of potential upcoming events with similar characteristics (4–6). Particularly complex are these issues in metropolitan, urban settings in which the dimensions of these complications are amplified as an effect of very large heterogeneous populations. Such was the case faced in Mexico City.

Mexico City, home to the highest concentration of tertiary care hospitals in Mexico, provides healthcare services not only to its 9 million permanent residents but also to an additional 13 million people from the surrounding metropolitan area and beyond. Despite this extensive healthcare infrastructure, the COVID-19 pandemic imposed unprecedented strain on the system, challenging its capacity to meet the rapidly growing demand for medical attention. In this regard, the pandemic exposed the (already existent, but possibly latent) inherent complexities of the health system, which operates as a multifaceted organization with numerous interdependent components. Traditionally viewed as hierarchical and linear, health systems are now recognized as complex adaptive systems characterized by non-linear interactions, self-organization, and emergent behavior (7). The sudden and unpredictable nature of the COVID-19 crisis (8) further amplified these complexities, revealing both the strengths and vulnerabilities of the healthcare network in Mexico City.

The present research systematically examines the evolving patterns of hospital demand during the pandemic, highlighting the dynamic shifts in patient behavior and hospital utilization. It was observed that, as the crisis intensified, individuals often sought care at hospitals far from their residences, resulting in uneven and entangled distribution of demand across the city’s healthcare facilities. While some hospitals experienced consistently high demand, others saw fluctuating levels of patient load, reflecting the adaptive and often chaotic nature of the pandemic response.

By analyzing these patterns, key factors were identified as contributors to the resilience of the health system, as well as areas where it struggled to cope with the extraordinary pressures of the pandemic. These insights may become crucial for developing strategies to enhance the system’s preparedness and adaptability in future health emergencies, ensuring a more effective and equitable response to unforeseen challenges.

## Results and discussion

### Hospital demand is a spatially dynamic variable that changes throughout the pandemic

The COVID-19 pandemic in Mexico City presented a complex and evolving landscape of hospital demand. Contrary to the assumption that certain areas would bear a consistent burden of cases, the spatial distribution of hospitalizations shifted notably over time, often in counterintuitive ways (9). This dynamic movement of cases reflects the heterogeneous patterns of exposure and vulnerability across different neighborhoods of the city, as well as structural biases in social development between boroughs and neighborhoods (10,11).

During the initial stages of the pandemic, cases were heavily concentrated in certain areas, primarily those with higher population density and relatively lower socioeconomic status (12). However, as the pandemic evolved, the geographical footprint of the virus expanded, reaching new regions drastically altering the patterns of hospital demand. This dispersion of cases is indicative of varied levels of exposure across the city, influenced by factors such as population mobility, compliance with public health measures, and local transmission dynamics (13).

To capture these shifts in hospital demand, the weighted centroid of hospitalizations over time was measured. This metric was useful to quantify the “center of mass” of hospital demand, weighted by the number of cases in each location. This analysis revealed a gradual but significant northward movement of the centroid as the pandemic progressed. Initially, demand was focused in the densely populated southern and eastern boroughs of Mexico City. Over time, however, this centroid shifted towards the northern regions, highlighting the spatially dynamic nature of the pandemic.

This northward drift in hospital demand underscores the adaptability of the healthcare system, which had to reallocate resources and adjust strategies in response to the changing geographical distribution of cases. Understanding these spatial dynamics is crucial for optimizing resource allocation and ensuring that all areas of the city have adequate access to medical care, particularly during periods of high demand.

**Figure 1:**
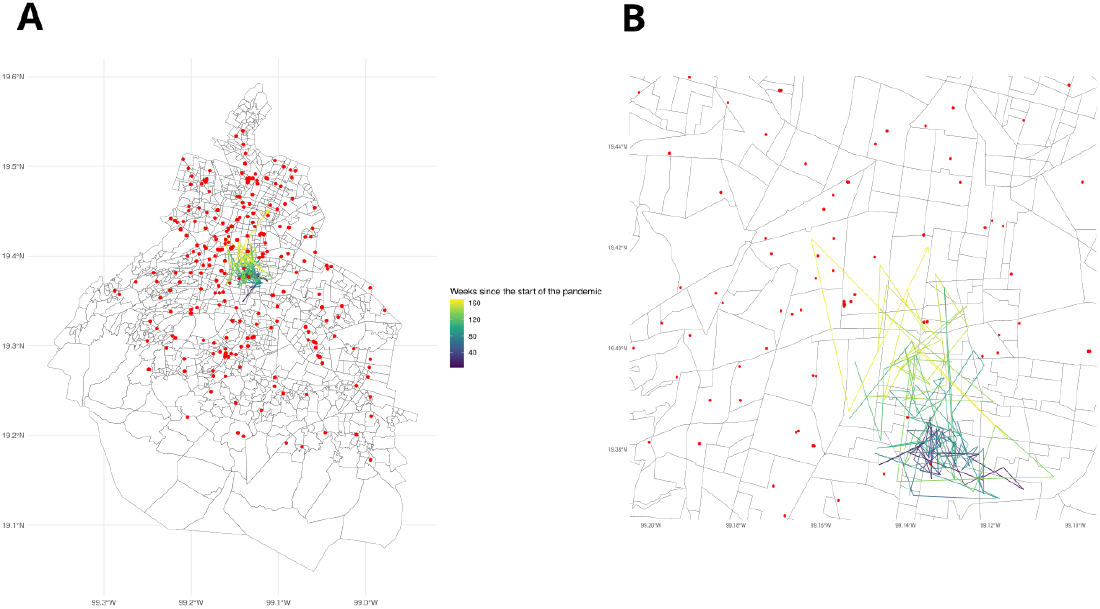
Map of Mexico City showing the trajectory of the hospitalization burden center of mass in terms of patient residence. Notice how this centroid shifts as the pandemic evolves. The location of hospitals habilitated for COVID-19 treatment is shown in red. Panel A: full map. Panel B: Zoom in to the center of mass trajectory.

### Nevertheless, hospitalizations were concentrated in a few hospitals throughout the pandemic

**Figure 2:**
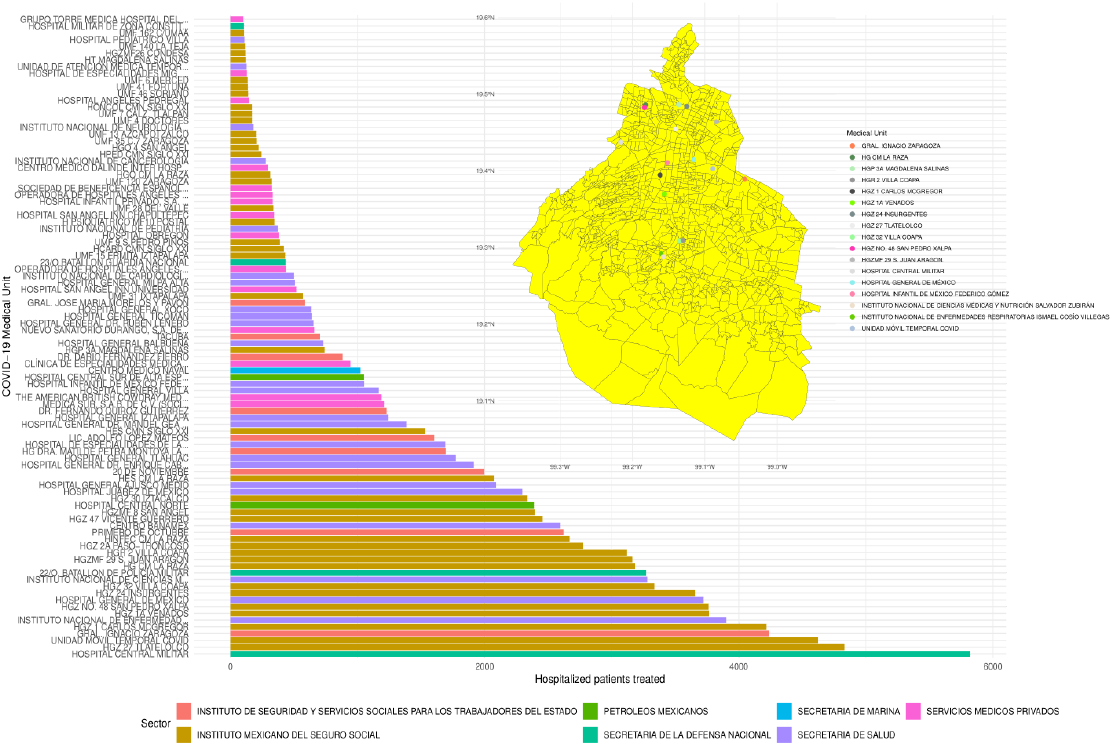
Barchart shows the distribution of cases treated by each COVID-19 hospital in Mexico City. Insert map shows the location of most consistently top ranked hospitals in terms of quantity of treated patients.

Despite the dynamic distribution of COVID-19 cases across Mexico City, the majority of hospitalizations were concentrated in a relatively small number of medical facilities. Specifically, 17 hospitals were ranked in the top 10 in terms of admissions for at least 35 weeks. These hospitals, spanning various sectors of the healthcare system—including the Mexican Social Security Institute (IMSS), the Institute for Social Security and Services for State Workers (ISSSTE), the Health Secretary, and the military—handled most of the hospital demand throughout the pandemic (9).

The consolidation of hospitalizations in these facilities reflects a multifactorial process shaped by population distribution, healthcare system structure, and resource allocation. Understanding how these hospitals came to dominate the demand landscape during different phases of the pandemic can offer valuable insights for optimizing resource distribution in future public health emergencies. For a population as socioeconomically and geographically disparate as Mexico City, comparing the evolution of these facilities’ demand with the locations of the most affected populations highlights key patterns in access to care and resource utilization.

A distinction exists between hospitals under the Federal Health Secretary, local healthcare services of Mexico City, and those belonging to IMSS and ISSSTE. IMSS and ISSSTE hospitals regularly serve enrolled beneficiaries, while federal and local hospitals absorbe a more heterogeneous patient population. Nevertheless, during the pandemic those institutions were instructed to provide services to the general population. However, there is no currently available data that quantitatively shows how this affected access to health services. Such data may lead to better insights on resource allocation.

**Figure 3:**
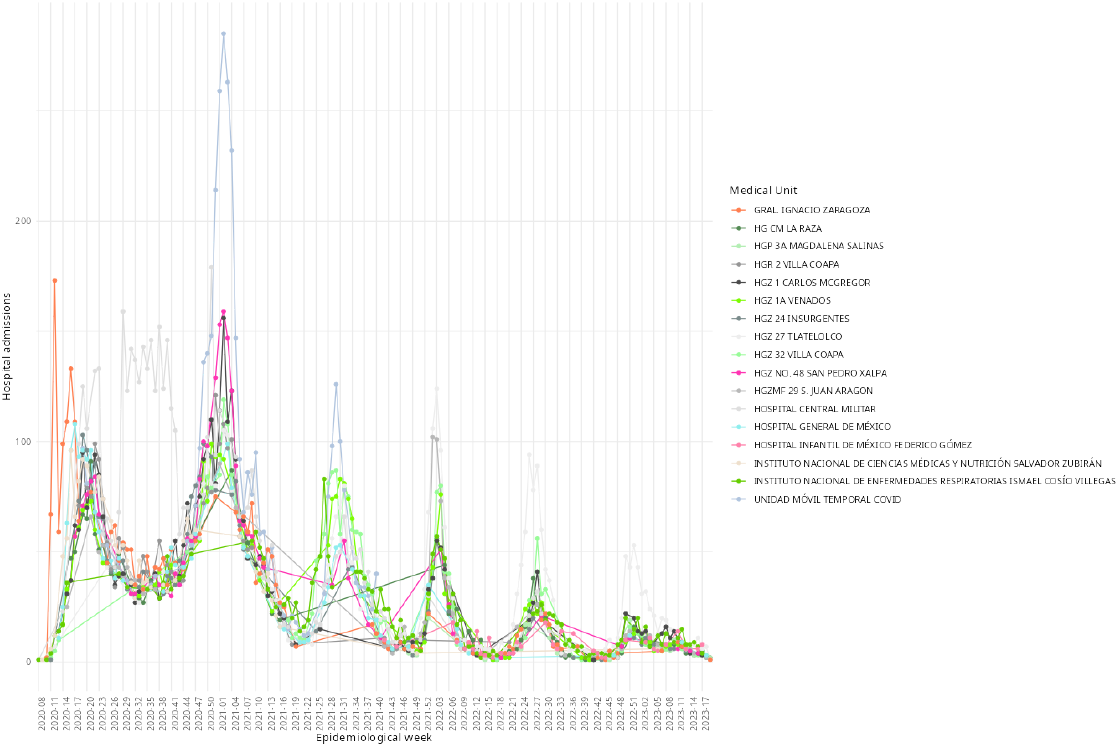
Time series of weekly hospitalizations in the most consistently top-ranked COVID-19 hospitals in Mexico City.

These hospitals experienced varying levels of demand over time, with most seeing significant increases during the high-contagion waves of the B.1.519 variant (14), the Delta variant, and, to a lesser extent, the first Omicron wave(15); demand waned, but did not disappear, as vaccination efforts advanced and reinfections came to dominate the epidemiological landscape (16). During the initial phase of the pandemic, the military hospital managed a substantial portion of the hospitalizations. In subsequent waves, the special COVID-19 temporal unit, established as a dedicated response facility in a large convention center in the northwestern part of the city, absorbed the majority of hospitalizations, particularly during the B.1.519 wave. This temporary unit represented an instance of adaptability within the otherwise static hospital infrastructure.

**Figure 4:**
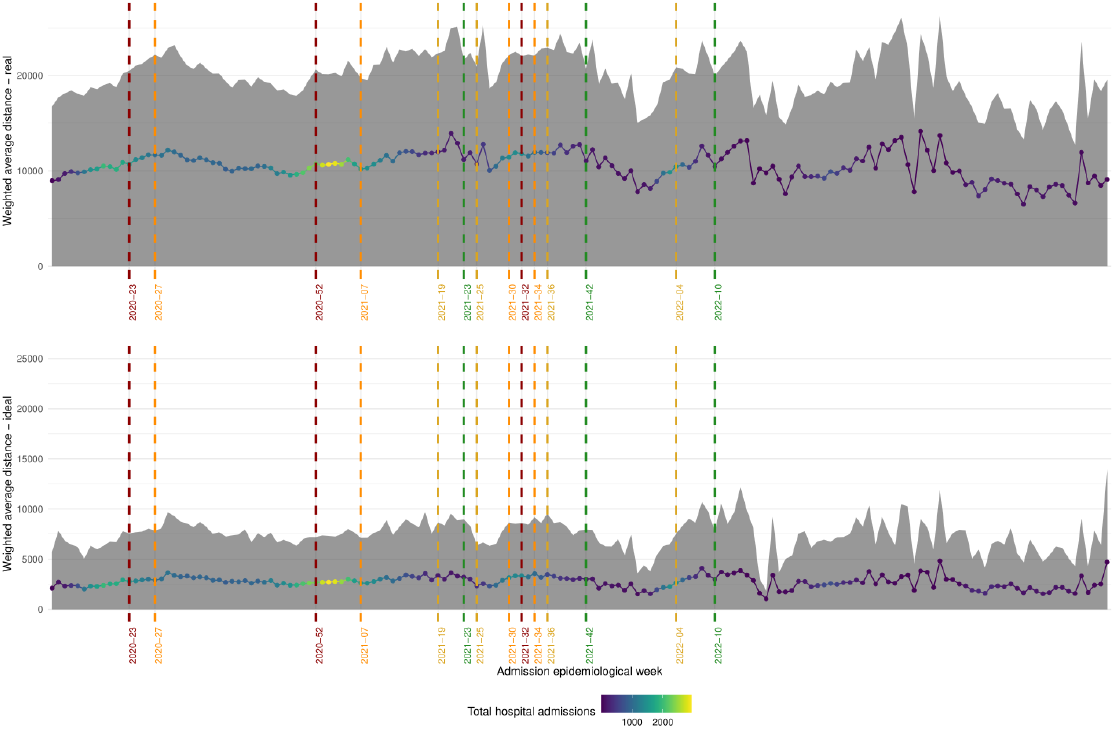
Time series of weighted average distance from home address to hospital for hospitalized patients of Covid-19. Dotted lines indicate transitions in the epidemiological stoplight system. Upper panel shows the empirical distances; lower panel shows the optimal distance (if patients were admitted to the clinic closest to their home).

### Hospital attention seeking behavior did not limit itself by spatial closeness

To understand the hospital attention-seeking behavior of COVID-19 patients in Mexico City, the distance patients traveled from their place of residence to the hospital where they received care was analyzed. The analysis focused on the weekly average distance traveled, calculated from the centroid of each patient’s ZIP code to the hospital where they were treated. This distance was then compared to an “optimal” scenario, in which patients would have been treated at the hospital closest to their residence.

The analysis revealed that the actual distance traveled by patients was nearly twice as long as the optimal distance throughout the pandemic. This discrepancy indicates that patients did not necessarily seek care at the nearest facility, even when geographical proximity would suggest otherwise. Instead, their hospital-seeking behavior was influenced by other factors, perhaps including perceived quality of care, hospital reputation, and possibly the availability of specialized services.

Moreover, the actual distance traveled by patients increased significantly during periods of high hospital saturation, such as during the peak waves of the pandemic, and following the relaxation of public health measures, such as the change in the epidemiological stoplight from red to orange. These patterns suggest that when hospital resources were strained or when restrictions were eased, patients were more likely to travel farther to seek care, possibly due to overcrowding at closer facilities or increased mobility.

The analysis of these travel patterns highlights the constraints precluding our healthcare system in evenly distributing hospital demand, as well as the behavioral flexibility of patients in seeking medical attention. It also underscores the need for strategies to reduce these travel burdens, such as improving resource allocation and communication to ensure that patients are aware of available care options closer to home, particularly during periods of high hospital demand.

It should be noted that hospital demand fluctuated in a closely correlated way for both general hospitalization and intensive care units (UCI). This is important as it highlights the level of stress and participation of each institution facing the pandemic. In general, it is expected that a higher demand of UCI beds would be indicative of a stronger impact on resource demands. As the figure shows, UCI demand scaled in considerably the same proportion throughout the first four waves. Only after vaccination efforts were widely deployed did the UCI demand decrease, as seen in the fifth wave.

**Figure 5:**
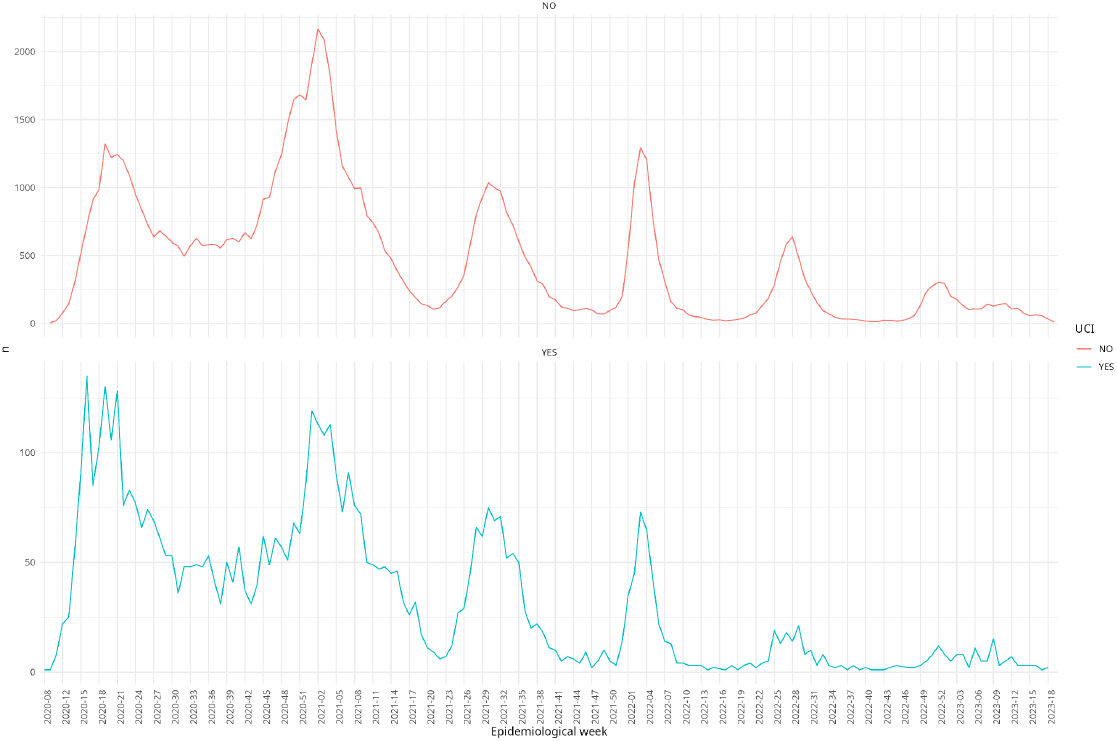
Comparison of hospital demand (measured in daily admissions) between general hospitalization beds and UCI beds.

### Higher hospital demand leads to phase transition in the hospital demand network

**Figure 6:**
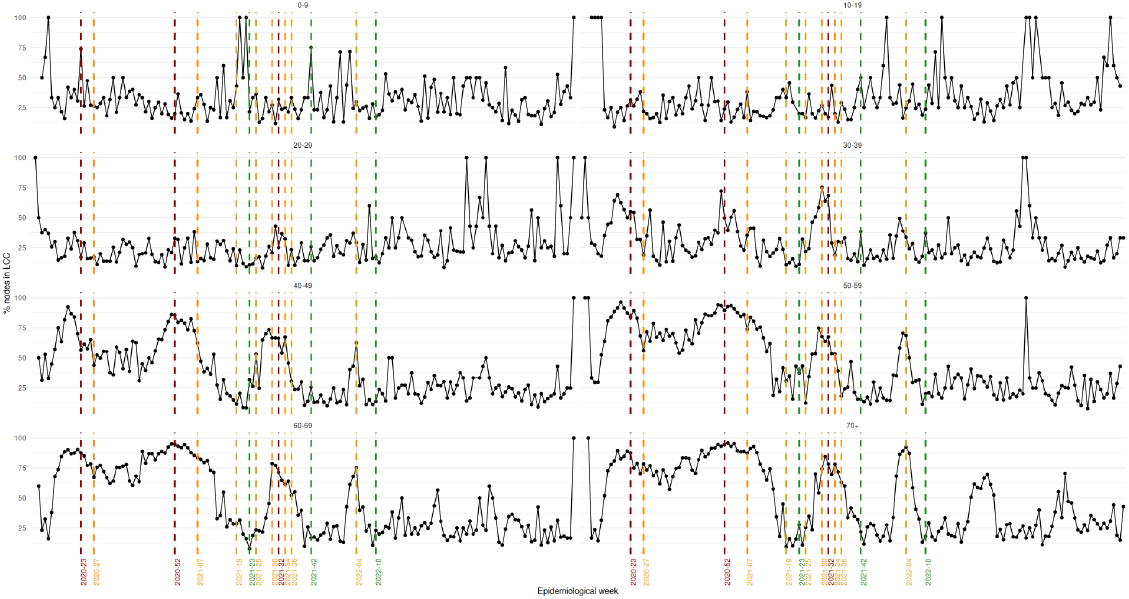
Time series of the size (in node percentage) of the largest connected component in the hospital demand network. Dotted lines indicate transitions in the epidemiological stoplight. Panels showcase different age groups.

To better understand the dynamics of hospital demand during the COVID-19 pandemic, the interaction between hospital demand and supply via a “hospital demand network” was modeled. This network is represented as a bipartite graph, where nodes are either residential areas (defined by ZIP codes) or hospitals, and connections are established when a hospital provides care to patients from a given area. The structure of this network evolves over time, reflecting changes in patient behavior and hospital utilization.

A key feature of this network is the emergence of giant components: connected subgraphs encompassing more than 50% of the nodes in the network. These giant components are indicative of a phase transition (17,18) in the hospital demand network, a phenomenon that occurs as the system transitions from a more ordered to a more disordered state (19) in response to increased demand.

During periods of lower hospital demand, the network is more structured and ordered, with patients predominantly seeking care at hospitals located closer (although not necessarily the nearest) to their place of residence. This results in a network composed of many small, distinct components, where a small set of hospitals serve a specific set of neighborhoods. However, as demand increases, this orderly structure breaks down. Patients from various areas begin to seek care at hospitals outside their immediate vicinity, either due to overcrowding or the unavailability of care at closer facilities.

This shift leads to the formation of a giant component in the network, characterized by hospitals admitting patients from a wide range of neighborhoods. The network becomes more disordered, reflecting the strain on the healthcare system and the loss of spatial constraints on patient behavior. This phase transition is a critical indicator of system overload and occurs just before or during periods of high hospital demand, such as during peak waves of the pandemic.

The emergence of giant components in the hospital demand network highlights the nonlinear dynamics of the healthcare system under stress. Understanding these transitions can provide valuable insights into the capacity limits of the system and the points at which patient care becomes increasingly disorganized. This knowledge is crucial for designing interventions aimed at mitigating the impact of surges in hospital demand and maintaining a more stable and effective healthcare network during future public health crises.

### And higher stress in the hospital network leads to higher risk for hospitalized patients

**Figure 7:**
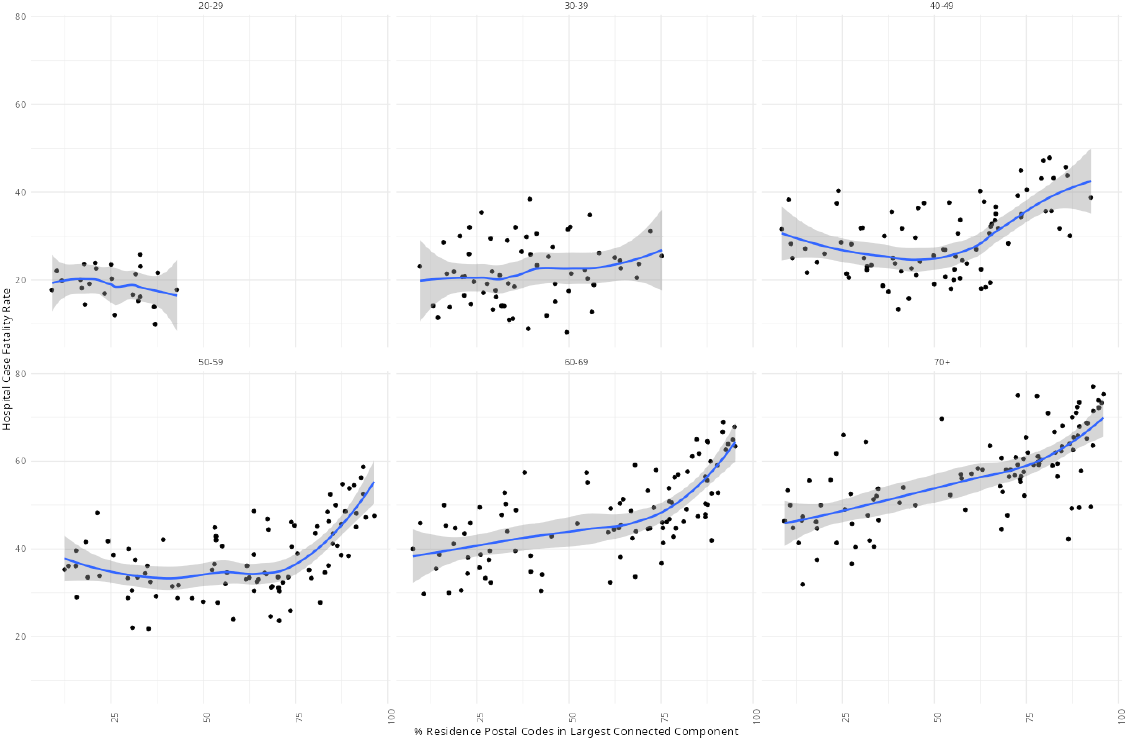
Scatterplot of hospital case fatality rate vs size of the largest connected component. Blue lines show Loess regression. Panels indicate different age groups.

Using the largest connected component (LCC) in the hospital demand network as a measure of “disordered strain” on the healthcare system, an increased strain—indicative of higher hospital demand—was associated with less favorable outcomes for hospitalized patients. As the LCC expands, encompassing a larger portion of the network, it reflects a breakdown in the structured distribution of patients across hospitals, suggesting that hospitals are increasingly overwhelmed and are treating patients from a broader range of neighborhoods.

This relationship between network strain and patient outcomes varied by age group. It was particularly pronounced among older patients, for whom age is already established as a primary risk factor for severe COVID-19 outcomes (20). In these older age groups, the correlation between increased network strain and adverse outcomes was not linear. Instead, it exhibited a phase transition: as the size of the giant component in the network grew, there was a marked increase in risk for hospitalized patients, particularly those over the age of 40. This is in agreement with what previous research had shown, particularly regarding non-comorbid patients (21).

This phase transition in risk suggests that the LCC, as a global measure of network strain, may be capturing the cumulative effects of increased pressure on individual hospitals. During periods of high demand, hospitals experience a higher patient load, potentially leading to resource shortages, longer wait times, and diminished quality of care—all of which can contribute to worse outcomes for patients as it was reported comparing temporary ICU’s units with conventional ICU’s (22) or poor availability of ICU beds as it was reported as well (23). The non-linear nature of this relationship underscores the complexity of the healthcare system, where local interactions and constraints can lead to emergent global phenomena that significantly impact patient care.

Understanding this dynamic is crucial for managing hospital resources and mitigating risk during periods of high demand. It highlights the need for interventions that not only address hospital capacity at a local level but also consider the broader network effects that can exacerbate strain across the healthcare system. Recognizing and responding to these critical phase transitions could be key to improving patient outcomes in future public health emergencies.

## Concluding remarks

The COVID-19 pandemic in Mexico City exemplifies the complex and dynamic nature of public health crises, characterized by multiple interacting layers of complexity. From the rapidly evolving epidemic dynamics to the intricate social behaviors and the challenges of resource allocation, the pandemic required a multifaceted response from the healthcare system. Adaptive measures, such as the establishment of temporary COVID-19 units, demonstrated the potential for effective, flexible responses to sudden surges in hospital demand. These temporary facilities, like the one set up in the northwestern part of the city, played a crucial role in managing the load on the healthcare system during critical periods, providing a blueprint for similar interventions in future emergencies.

The success of such adaptive strategies highlights the importance of developing and refining these approaches for rapid deployment in response to evolving public health challenges. Enhancing the capacity to implement temporary medical facilities, along with other flexible resource allocation strategies, can significantly improve the system’s resilience to unexpected demand surges.

Furthermore, the application of measures of phenomenological complexity, such as network analysis of hospital demand, can provide valuable early warning indicators. These tools can help identify critical phase transitions and emerging strains on the healthcare system, enabling just-in-time decision-making and rapid response. Incorporating these complex system metrics into public health planning and response strategies will be essential for anticipating and mitigating the impact of future health emergencies.

## Methods

### Epidemiological Data

For this study, data from the centralized database for COVID-19 case reporting system (SISVER) was used. SISVER (respiratory disease epidemiological surveillance system) was managed by the Mexican federal government. The dataset is a “line-list” format, with each row representing an individual case. It is important to note that all information in this database is collected exclusively from official public reporting institutions at the state or federal level.

As part of the Federal Health System, our institutions were granted access to the complete dataset, comprising 130 variables per case. For this analysis, the study focused on key variables relevant to the research objectives, including the patient’s ZIP code of residence, date of symptom onset, age, hospitalization status, and the medical unit where the patient received care.

This dataset contained detailed insights into the spatiotemporal patterns and dynamics of COVID-19 hospitalizations within the study population. Only cases meeting the following criteria were included: (i) confirmed COVID-19 cases, verified through PCR or antigen testing; (ii) cases recorded in medical units or testing sites located within Mexico City boundaries; (iii) patients with a reported residence in Mexico City; and (iv) cases collected up to epidemiological week 16 of 2022 that required hospitalization. This selection resulted in a total of 122,933 cases analyzed.

The study was approved by the ethics and research committees of the Instituto Nacional de Medicina Genómica (approval codes CEI/1479/20 and CEI 2020/21).

### Medical Unit Data

Data on medical units were obtained from the Catalog of Unique Keys for Medical Units (CLUES), a federal database providing information on medical units across Mexico. Using the geolocation data provided in this catalog, the road distance between each medical unit and the centroid of each ZIP code in Mexico City was calculated.

### Evaluation of Distance Traveled for COVID-19 Medical Attention

The travel behavior of patients requiring hospitalization was obtained by calculating the distance they traveled from their place of residence (approximated by the centroid of their ZIP code) to the medical unit where they received treatment. The weekly weighted average distance traveled was computed using the formula:

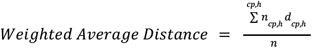

Where *n*_*cp*,*h*_ is the number of patients from ZIP code *cp* in hospital *h*, d_cp,h_ is the distance between the (centroid of) ZIP code *cp* and hospital *h*, and n is the total number of patients requiring hospitalization in a given week. We compared this empirical distance to an *ideal* distance scenario, in which all patients received care at the medical unit closest to their residence.

#### Hospital Territorial Demand Network Model

To capture the complexity of hospital demand dynamics throughout the pandemic, a series of weighted bipartite networks representing the relationships between ZIP codes (residential areas) and hospitals were constructed. For each week, a network with ZIP codes *cp* and hospitals *h* as nodes, connected by edges when at least one resident of ZIP code *cp* was treated at hospital *h* was created. The weight of each edge represented the number of patients from that ZIP code treated at the hospital.

This analysis focused on the largest connected component (LCC) of the network, defined as a component containing more than half of the nodes. The emergence of giant components over time, which indicate a phase transition in the network structure, was monitored. These transitions reflect changes in hospital demand patterns, especially during the progression of the pandemic and hospital system strain.

## Data Availability

All data produced in the present study are available upon reasonable request to the authors

## Notes

### Competing Interest Statement

The authors have declared no competing interest.

